# AI is Smart. Is it Wise? Quantifying the Effect of Patient-Choice (β) on Physical Outcomes^*^

**DOI:** 10.64898/2026.03.10.26348069

**Authors:** Ogan Gurel, Matthew F. Rasmussen, Venkatesh Veginati, James N. Weinstein

## Abstract

Large language models (LLMs) increasingly guide clinical decisions through population-level evidence, yet they cannot encode individual patient preferences. When treatments yield comparable outcomes, patient choice may drive decisions, though its effect remains unquantified. The Spine Patient Outcomes Research Trial (SPORT)—marked by similar surgical and nonoperative results and substantial crossover—provided a natural experiment to apply causal-inference methods to estimate unbiased treatment effects and quantify the contribution of patient choice to outcomes. Using only published aggregate results from SPORT, we conducted two-stage least squares instrumental-variable analysis with randomized treatment assignment as the instrument, with Complier Average Causal Effects (CACE) and E-values assessing sensitivity to unmeasured confounding. Primary outcomes were SF-36 Bodily Pain, SF-36 Physical Function, and the Oswestry Disability Index. We decomposed treatment effects into **α**, the biological treatment mechanism, and **β**, the preference-driven selection component reflecting the downstream effect of patient choice. Aggregate estimates revealed **α**_G_ = 15.7 (0.5) and **β**_G_ = 7.4 (3.4), with a negligible net difference between surgical and nonoperative treatment effects (Δ**α** ≈ 0.65). This analysis demonstrates that patient choice (**β**) is a measurable and clinically meaningful contributor to physical outcomes. When treatment effects are comparable, **β**—a dimension inaccessible to current LLMs trained primarily on **α**-biased population-level evidence—emerges as the dominant driver of decision-making. These findings provide empirical grounding for informed choice, clarify the limits of present-day AI in preference-sensitive care, and identify a structural constraint in AI-driven clinical decision support.

**Key messages:**

- The effect of patient choice (**β**) on physical outcomes is real, measurable, and clinically meaningful.
- **β** becomes the dominant driver of outcomes when biological treatment differences (Δ **α**) are small.
- LLMs cannot encode **β** because they are trained on α-biased population-level evidence.
- These findings provide the empirical foundation for informed choice—not just informed consent.

## Introduction

Artificial-intelligence systems are increasingly embedded in clinical medicine, supporting diagnosis, guiding treatment, and informing prognosis.^1,2,3,4^ When evidence-based guidelines are equivocal, however, large-language-model systems trained on those datasets cannot offer decisive recommendations. As Dr. John Wennberg noted, “preference-neutral” conditions— where evidence identifies a clearly superior treatment—contrast with “preference-sensitive” conditions, in which multiple reasonable options differ in benefits, risks, and the values patients assign to them.^5,6^ The former lies squarely within evidence-based medicine (EBM), whereas the latter requires values-based medicine (VBM). AI systems perform well in the former domain but remain fundamentally limited in the latter.^7^

The Spine Patient Outcomes Research Trial, SPORT, exemplifies this paradigm. When comparing surgery with nonoperative treatment for lumbar disc herniation, SPORT demonstrated clinical equipoise, with both approaches yielding substantial improvements and overlapping confidence intervals, although 42–45% of randomized participants crossed over to the alternative treatment within 3 months.^8,9,10^ Traditional interpretation treats such crossover as a protocol violation that dilutes intention-to-treat (ITT) estimates.^11,12^ We considered an alternative: high crossover is signal, not noise—evidence of strong, directionally consistent, and outcome-shaping patient preference.

This reframing sharpens the distinction between informed consent and informed choice. Informed consent protects patient autonomy in preference-neutral conditions in which evidence and clinical expertise identify a clearly superior treatment and allow LLM-based decision support to contribute meaningfully. Preference-sensitive conditions, however, require informed choice— a deliberative process that aligns benefit–harm profiles with patient values through explicit preference elicitation.

This distinction becomes critical for AI deployment. Although AI systems can predict outcomes from population-level data, they cannot represent the value-dependent trade-offs that guide individual treatment decisions. LLMs may summarize the evidence supporting different treatment strategies, but they cannot discern how an individual patient weighs clinical risks, treatment burden, adherence demands, medication side effects, pain tolerance, financial considerations, or competing life priorities.^13,14^

Recent systematic evaluations reveal fundamental architectural constraints in current LLMs when applied to preference-sensitive care. A recent scoping review found that while AI excels at processing population-level data, such systems lack validation in real-world settings in which preference-sensitive decisions are made.^15^ Similarly, a study demonstrated that LLMs fail to follow instructions and are sensitive to both quantity and order of information, performing significantly worse than clinicians when clinical decision-making requires integrating patient-specific utilities.^16^ The limitation is architectural: LLMs learn conditional outcome probabilities from evidence-based datasets but cannot represent the individual utility functions that drive value-dependent decisions. When treatment effects, which we call **α**, are comparable, the optimal choice depends on patient-specific value functions—precisely the domain where token-based language models, lacking access to real-time preference elicitation, cannot operate. While LLMs summarize evidence, they lack knowledge of the patient-specific utility function that determines whether surgical risk is acceptable for a given individual.^17^ That determination depends on what we term **β**, a parameter fundamentally inaccessible to systems trained purely on historical outcomes that are generally biased to tabulate **α** effects. These limitations have implications not only for clinical decision support but also for regulatory frameworks, reimbursement models, and the ethical deployment of AI in patient-centered care.

We applied causal-inference methods to estimate treatment effects under crossover^18,19^ and to quantify selection bias as patient choice. This included assessing whether the patient-choice parameter (**β**) is significant, how it compares with the treatment-effect parameter (**α**), and how **β** relates to the treatment-effect difference when **α**_surgery_ and **α**_nonoperative_ are similar. Evidence-based medicine and values-based medicine function as complementary domains of clinical decision-making. In contrast to prior work, which has been conceptual or normative^18,19^ we provide causal-inference evidence that patient choice is significant, measurable, and clinically meaningful—thereby offering empirical support for VBM. Finally, we examined how these findings challenge the capacity of current AI systems to guide care in preference-sensitive settings, where wisdom—not merely predictive accuracy—requires incorporating the influence of patient choice.

## Methods

### STUDY DESIGN

We conducted a secondary analysis of published SPORT data comparing surgical discectomy with nonoperative treatment for lumbar disc herniation^8,9^. Eligible participants had radicular pain with imaging-confirmed herniation and symptoms lasting at least six weeks. Patients choosing their treatment enrolled in the observational cohort (n=743), whereas those willing to be randomized were assigned to surgery (n=245) or non-operative care (n=256). Primary outcomes were SF-36 Bodily Pain (BP), SF-36 Physical Function (PF), and the Oswestry Disability Index (ODI) measured at 3, 12, and 24 months^20^ Minimal clinically important differences were 4–5 points for the SF-36 scales and 8–10 points for the ODI.^21,22,23,24^

### STATISTICAL ANALYSIS

#### Intention-to-Treat Analysis: Estimation of Treatment Effects (α)

Randomized groups were compared by assignment to obtain intention-to-treat (ITT) estimates, which preserve randomization but attenuate effect sizes under crossover. The ITT treatment effect was defined as **α**, with **α**_G_ representing the average ITT effect across time points.

#### Causal-Inference Estimation (IV and CACE)

Our primary analytic approach used instrumental-variable (IV) methods, with randomized treatment assignment serving as the instrument for treatment received.^25,26^ Causal estimates were obtained under standard IV assumptions—relevance, exclusion, independence, and monotonicity. We implemented two-stage least squares (2SLS), modeling treatment received as a function of assignment and baseline covariates in Stage 1, and outcomes as a function of predicted treatment in Stage 2; a first-stage *F*-statistic greater than 10 indicated instrument strength. We calculated the Complier Average Causal Effect (CACE) using two approaches: CACE*1*, the ITT effect divided by the compliance rate, and CACE*2*, the 2SLS-based estimator aligned with the IV framework.

#### Estimation of Treatment Effects (α_surgery_, α_nonoperative_, and α_G_)

We constructed a direction-consistent composite treatment effect at each time point by reversing ODI and averaging it with BP and PF, placing all measures on a 0–100 scale (higher=better). This composite corresponds to the randomized contrast Δ_ITT_ and is denoted **α**. Overall **α**_surgery_ and **α**_nonoperative_ effects were defined as the average of these values across time points. Finally, a treatment-agnostic summary, **α**_G_, was calculated as the mean of **α**_surgery_ and **α**_nonoperative_.

#### Estimation of the Patient-Choice Effect (β and β_G_)

With **α** representing the randomized contrast Δ_ITT_, we defined its observational analogue, Δ_OBS_. The divergence between Δ_OBS_ and Δ_ITT_ isolates variation attributable to patient choice, with this residual difference quantifying the influence of preference-driven selection on outcomes. We call this **β** and quantified it using four estimators, ordered by increasing causal rigor:

1. **β**_Arithmetic;_=Δ_OBS;_−Δ_ITT_
2. **β**_CACE1_=Δ_OBS_–Δ_CACE*1*;_ (ITT/π)
3. **β**_CACE2_=Δ_OBS;_–Δ_CACE*2*_ (2SLS)
4. **β**_IV_=Δ_OBS_–Δ_IV_

To estimate an aggregate patient choice effect, we averaged across outcomes, estimators, and time points to obtain **β**_G_.

#### Interpreting β as preference-driven selection

By construction, **β** is defined as the divergence between observational contrasts and causal estimates under randomized assignment. This residual captures outcome variation arising from patient selection, not from biological treatment differences. Factors such as expectations, motivation, adherence, or risk tolerance may mediate outcomes downstream, but they operate through the act of choice itself. Within the causal-inference framework, these mechanisms are not alternatives to preference-driven selection but components of it. The convergence of IV and CACE estimates, their persistence across outcomes and time, and robustness to unmeasured confounding (E-values) support interpreting **β** as a structured, preference-mediated causal component rather than undifferentiated behavioral noise.

#### Sensitivity Analyses (*E*-values and Bootstrap)

To assess robustness to unmeasured confounding, we calculated *E*-values, which quantify the minimum confounder strength needed with both treatment and outcome to explain the observed effect. We also generated 10,000 bootstrap resamples to estimate 95% confidence intervals. All analyses followed CONSORT guidelines for trials with non-compliance. Two-sided P<0.05 was considered significant.

## Results

### STUDY POPULATION AND INSTRUMENT VALIDATION

SPORT enrolled 1,244 participants (randomized cohort, n=501; observational cohort, n=743). At 3 months, 42–45% of randomized participants crossed over, consistent with strong patient preference. The first-stage regression confirmed instrument strength (*F*=87.3; partial *R*^2^=0.43; P < 0.001). Cumulative compliance differences (π) were 0.200 (0.044) at 3 months, 0.166 (0.045) at 12 months, and 0.158 (0.045) at 24 months, supporting relevance and monotonicity.

### TREATMENT EFFECTS (α)

#### SF-36 Bodily Pain (BP)

At 3 months, the intention-to-treat (ITT) comparison showed a diluted effect (2.9 points, 95% CI −0.9 to 6.7). Causal estimators were larger: IV was 8.7 (95% CI 3.8 to 13.6) and CACE 9.2 (95% CI 4.1 to 14.3). The observational contrast (patients selecting treatment) was 14.9 (95% CI 10.9 to 18.9), exceeding causal estimates by approximately 6–7 points (∼71% inflation).

#### SF-36 Physical Function (PF)

ITT showed −0.5 (95% CI −4.3 to 3.3); causal estimators were IV 9.2 (95% CI 4.5 to 13.9) and CACE 9.8 (95% CI 4.9 to 14.7). The observational contrast was 15.4 (95% CI 11.6 to 19.2), again approximately 6 points above causal estimates (∼ 67% inflation).

#### Oswestry Disability Index (ODI)

ITT was −4.7 (95% CI −8.5 to −0.9); causal estimators were IV −13.1 (95% CI −18.2 to −8.0) and CACE −13.9 (95% CI −19.2 to −8.6). The observational contrast was −15.2 (95% CI −18.5 to −11.8), just two points beyond causal estimates (∼16%). Across outcomes at 3 months, all IV/CACE effects exceeded minimal clinically important differences, whereas ITT estimates were attenuated by crossover.

#### Treatment Effects Persisted

At 12 months, IV estimates were 7.8 (BP), 8.6 (PF), and −11.4 (ODI); at 24 months, 7.1 (BP), 8.2 (PF), and −10.8 (ODI). With bootstrap confidence intervals applied to IV and CACE estimates at 12 and 24 months, patterns remained concordant. Uncertainty intervals widened, but results continued to favor clinically meaningful benefit among compliers. Observational contrasts continued to overestimate effects relative to IV/CACE by approximately 5–7 points for SF-36 domains and approximately 3 points for ODI.

#### Analytical Concordance

IV (8.7) and CACE (9.2) estimates closely agreed—two distinct causal inference converged (Figure 1). Patterns of ITT underestimation, IV/CACE convergence, and observational overestimation were consistent.

**Figure 1.**
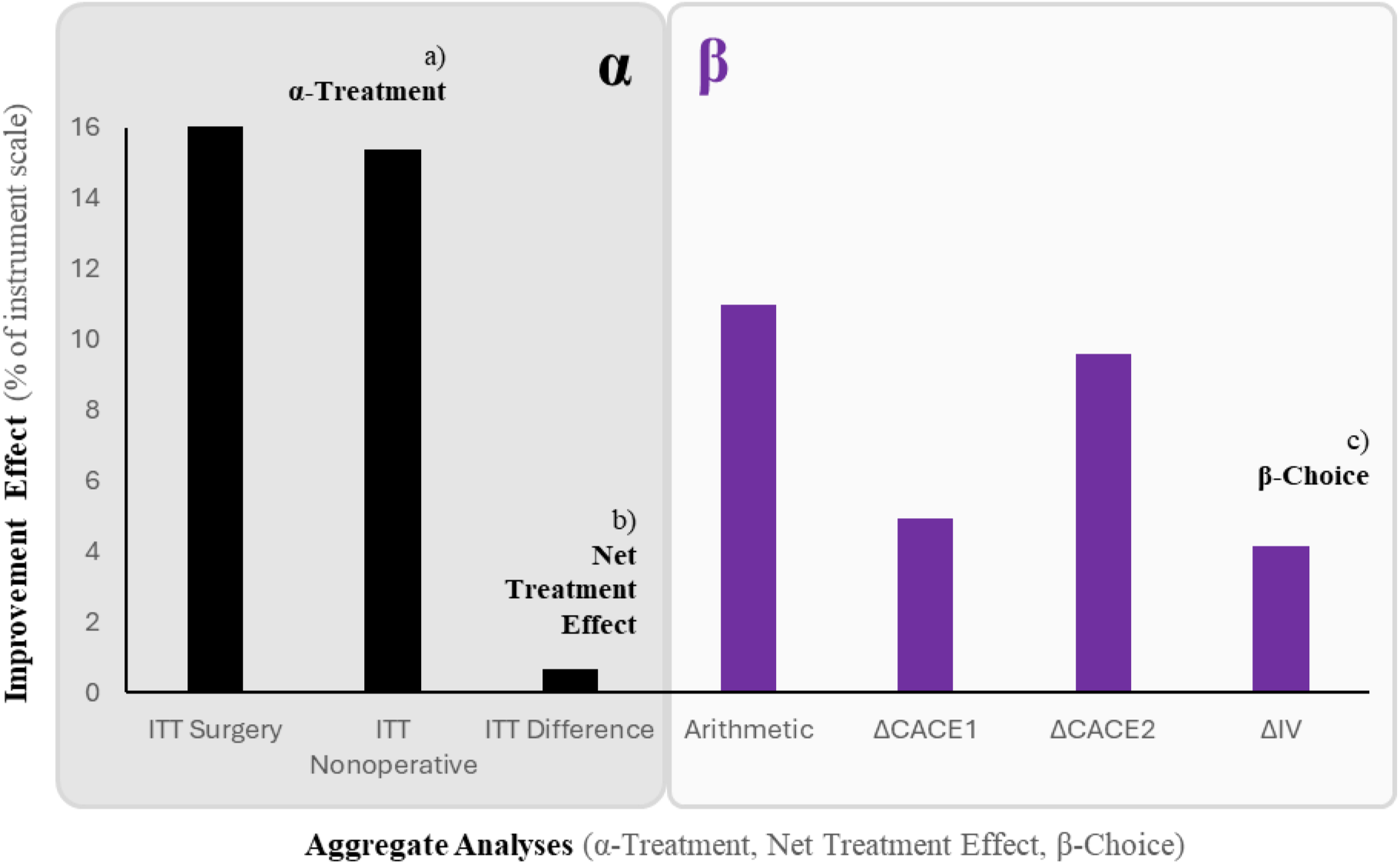
Aggregate Treatment and Choice Effects Across Analytic Approaches Shown are the aggregated estimates, **α**_G_, for (a) the **α**_Treatment effect_, (b) the Net Treatment Effect (**α**_surgery_ **- α**_nonoperative_), and (c) the **β**_Choice_ effect calculated using four predefined methods (Δ_Arithmetic_, Δ_CACE*2*_, Δ_CACE*1*_, and Δ_IV_). The **α**_Treatment_ panel displays both ITT_Surgery_ and ITT_Nonoperative_ having a favorable effect on treatment while Net Treatment Effect panel displays the difference, showing the two approaches to be equivalent; the **β**_Choice_ panel displays the magnitude for each **β** estimator.

### PATIENT CHOICE EFFECT (β)

As mentioned, the **β**-choice effect was calculated using four methods: **β**_Arithmetic_, **β**_CACE1_, **β**_CACE2_, and **β**_IV_, as summarized in Table 3. At 3 months, Δ_OBS_ exceeded Δ_ITT_ by 12.0 points for BP, 15.9 points for PF, and 10.5 points for |ODI|, with consistent but smaller residuals when contrasted against CACE/IV. The aggregate patient choice impact, **β**_G_, was 7.4 (3.4) points. Formal Z-tests demonstrated significant choice-associated divergence across outcomes and time. At 3 months, for example, BP z = 3.57 (P=3.6×10^−4^), PF z = 3.78 (P=1.6×10^−4^), and ODI z = 3.61 (P=3.1×10^−4^), with comparable results at 12 and 24 months.

**Table 1.** Baseline Characteristics and Crossover Patterns.

| **Table 1. Baseline Characteristics and Crossover Patterns** |  |  |  |  |
| --- | --- | --- | --- | --- |
|  | **Randomized**<br>**Surgery**<br>(n = 245) | **Randomized**<br>**Nonoperative**<br>(n = 256) | **Observational**<br>**Surgery**<br>(n = 526) | **Observational**<br>**Nonoperative**<br>(n = 217) |
| **Age**<br>mean (SD), years | 41.7 (11.2) | 42.3 (10.8) | 40.8 (10.9) | 43.1 (11.5) |
| **Female**<br>% | 40.0 | 41.4 | 41.4 | 43.8 |
| **ODI**<br>mean (SD) | 48.2 (16.3) | 47.8 (17.1) | 49.6 (15.8)* | 46.8 (17.2)* |
| **SF-36 Pain**<br>mean (SD) | 28.4 (18.7) | 29.1 (19.2) | 27.8 (18.3) | 30.2 (19.5) |
| **Crossover**<br>by 3 months, % | 42.0 | 41.8 | — | — |
| **Compliance**<br>**rate**<br>% | 58.0 | 58.2 | — | — |
| *P=0.02 for ODI difference between observational groups. Substantial crossover in randomized cohort validates preference strength. |  |  |  |  |

Baseline demographic and clinical characteristics were similar across randomized and observational cohorts. ODI differed between observational groups ( $P = 0.02$ ). High crossover rates in the randomized cohort (42–45% by 3 months) reflect strong patient preferences and validate the relevance of treatment assignment as an instrument.

**Table 2.** Treatment Effects Across Analytic Approaches.

| Outcome | Time<br>(months) | ITT | IV | CACE | Observational | IV<br>E-value<br>(point) | IV<br>E-value<br>(CI-limit) |
| --- | --- | --- | --- | --- | --- | --- | --- |
| **BP** | **3** | 2.9<br>(-0.83, 6.63) | 8.7<br>(3.8, 13.6) | 9.2<br>(4.1, 14.3) | 14.9<br>(10.77, 19.03) | 1.9 | 1.4 |
|  | **12** | 2.8<br>(-0.74, 6.34) | 7.8<br>(-30.1, 46.6) | 8.3<br>(-29.6, 47.1) | 10.8<br>(6.50, 15.10) | 1.8 | 1.3 |
|  | **24** | 3.2<br>(-0.53, 6.93) | 7.1<br>(-38.1, 48.4) | 7.6<br>(-37.6, 48.2) | 10.2<br>(5.90, 14.50) | 1.7 | 1.3 |
| **PF** | **3** | -0.5<br>(-5.45, 4.45) | 9.2<br>(4.5, 13.9) | 9.8<br>(4.9, 14.7) | 15.4<br>(11.53, 19.27) | 2 | 1.5 |
|  | **12** | 1.2<br>(-3.75, 6.15) | 8.6<br>(-30.0, 46.5) | 9.1<br>(-29.5, 22.2) | 15.1<br>(10.90, 19.30) | 1.9 | 1.4 |
|  | **24** | 0.0<br>(-5.10, 5.10) | 8.2<br>(-31.1, 52.4) | 8.7<br>(point only) | 12.0<br>(7.80, 16.20) | 1.8 | 1.4 |
| **ODI** | **3** | -4.7<br>(-8.94, -0.46) | -13.1<br>(-18.2, -8.0) | -13.9<br>(-19.2, -8.6) | -15.2<br>(-18.60, -11.80) | 2.2 | 1.7 |
|  | **12** | -3.2<br>(-7.44, 1.04) | -11.4<br>(-47.4, 22.9) | -12.1<br>(-49.1, 22.2) | -15.3<br>(-19.03, -11.57) | 2 | 1.6 |
|  | **24** | -2.7<br>(-6.94, 1.54) | -10.8<br>(-47.3, 29.5) | -11.5<br>(point only) | -13.4<br>(-17.13, -9.67) | 1.9 | 1.5 |

Treatment effects for SF-36 Bodily Pain, Physical Function, and the Oswestry Disability Index at 3, 12, and 24 months are shown across intention-to-treat, instrumental-variable, and CACE estimators, with corresponding observational contrasts. IV E-values reflect the minimum strength of unmeasured confounding required to explain the causal estimates.

**Table 3.** β-Choice Estimates Calculated Using Four Methods.

| **Outcome** | **Time**<br>**(months)** | **$\beta_{\text{Arithmetic}}$** | **$\beta_{\text{CACE1}}$** | **$\beta_{\text{CACE2}}$** | **$\beta_{\text{IV}}$** |
| --- | --- | --- | --- | --- | --- |
| **BP** | **3** | 12 | 5.7 | 6.2 | 5.8 |
|  | **12** | 8 | -6.102 | 2.5 | 3 |
|  | **24** | 7 | 2.956 | 2.6 | 3.1 |
| **PF** | **3** | 15.9 | 1.39 | 5.6 | 6.2 |
|  | **12** | 13.9 | -5.303 | 5.9 | 6.4 |
|  | **24** | 12 | 3.3 | 3.8 | 8.2 |
| **ODI** | **3** | 10.5 | 8.317 | -1.3 | -2.1 |
|  | **12** | 12.1 | 4.117 | -3.1 | -3.8 |
|  | **24** | 10.7 | 3.73 | -1.9 | -2.6 |

β-Choice values were calculated as the difference between the observational contrast and each of four analytic estimators: $\beta_{\text{Arithmetic}} = \Delta_{\text{OBS}} - \Delta_{\text{ITT}}$ , $\beta_{\text{CACE1}} = \Delta_{\text{OBS}} - \Delta_{\text{ITT}/\pi}$ , $\beta_{\text{CACE2}} = \Delta_{\text{OBS}} - \Delta_{\text{CACE}(2\text{SLS})}$ , and $\beta_{\text{IV}} = \Delta_{\text{OBS}} - \Delta_{\text{IV}}$ . Values are reported for BP, PF, and the ODI at 3, 12, and 24 months.

**Table 4.** Key Findings.

| Table 4. Key Findings |  |  |  |
| --- | --- | --- | --- |
| Finding | Statistical Evidence | Clinical Interpretation | AI Implication |
| **$\beta$ is significant** | $Z = 3.57-3.78$,<br>$P < 0.001$ across outcomes | Patient choice contributes measurable effect | LLMs miss this parameter entirely trained only on outcomes, not preferences |
| **$\beta$ is clinically meaningful** | $\beta_G = 7.4$ (3.4) points; exceeds MCID | Effect size comparable to many interventions | AI recommendations ignoring $\beta$ may mis-rank options |
| **$\beta > \alpha$ difference in equipoise** | Net $\alpha$ difference = 0.65 pts;<br>$\beta_G = 7.4$ pts (11× larger) | Patient values determine choice when treatments similar | Precisely where LLMs least useful—cannot differentiate |
| **Crossover is signal** | 42-45% crossover; IV estimates 3 times ITT | Patients act on preferences despite randomization | AI trained on ITT underestimates effects; observational data overestimates 71% |
| **$\beta$ persists longitudinally** | Significant at 3, 12, 24 months | Preference effects influence long-term trajectories | AI systems must account for preference-driven selection throughout care |
| **Robustness confirmed** | $E$-values 1.7-2.2 | Unmeasured confounding unlikely to explain $\beta$ | Evidence that $\beta$ represents true causal effect |

This table compares analytic methods by their ability to support causal inference, handle treatment crossover, address selection bias, and align with NEJM reporting standards. It clarifies each method's role in estimating treatment effects and quantifying bias.

#### Robustness to unmeasured confounding

*E*-values ranged from 1.7–2.2 for point estimates and 1.3–1.7 for CI limits, indicating that an unmeasured confounder would need associations of that magnitude with both treatment and outcome to explain the effect.

### COMPARISON OF α AND β

In aggregate analyses (Figure 1), the choice effect (**β**) was consistently positive across all four predefined estimators (panel c), with values ranging from 4.1 to 10.9 points and an average **β**_G_ of 7.4 (3.4) points. Although **β**_G_ was smaller than the overall treatment effect **α**_G_ (15.7 [0.5]), it exceeded the net difference between surgical and nonoperative **α** estimates, which was negligible at 0.65 points (panel b).

## Discussion

### PRINCIPAL FINDINGS

In this secondary analysis of SPORT, treatment-effect estimates varied by analytic approach in the presence of substantial crossover, with IV and CACE yielding larger early effects than ITT and differences that persisted at 12 and 24 months. These patterns held across Bodily Pain, Physical Function, and the Oswestry Disability Index, and the instrument met accepted first-stage strength criteria. From these data, we addressed three questions: (1) whether patient choice (**β)** is significant; (2) how **β** compares with the treatment effect (**α**); and (3) how this comparison behaves when Δ**α** across treatments is minimal. Taken together, these findings show that patient choice is a causal contributor to physical outcomes, clarify its role in clinical decision-making, and delineate the limits of AI in preference-sensitive care.

### β > 0: THE PATIENT CHOICE EFFECT IS REAL

We quantified the contribution of patient choice using four estimators—**β**_Arithmetic_, **β**_CACE*1*_, **β**_CACE*2*_, and **β**_IV_ —each defined as the residual difference between observational and causal contrasts. Across outcomes and time points, all yielded positive values, with a simple mean **β**_G_ of 7.4 (3.4) points, demonstrating that **β** ≠ 0—a *non-vanishing* quantity. Its magnitude was clinically meaningful: **β** estimates were within or above established MCID thresholds for the SF-36 domains and the ODI. Statistical support came from Z-tests and a design-level meta-regression. The concordance of IV and CACE estimates, together with the systematic elevation of observational contrasts over ITT, reinforced internal consistency. These findings affirm that patient choice contributes a measurable and causally identifiable component to physical outcomes.

### β < α_G_: TREATMENT OUTWEIGHS PATIENT CHOICE

Because treatment effects anchor clinical decision-making, the aggregate treatment effect (**α**_G_) would be expected to exceed that of choice. In our analysis, **β** was indeed smaller than **α**_G_ yet remained measurable across methods, showing that patient choice shapes outcomes even as treatment effects remain larger. At first glance, **β** might seem *covariate* or even noise; in truth, it constitutes a *systematic causal component*.

This study has limitations. Our estimates of **β** may understate the true effect of patient preference, as some non-crossover patients may have made passive choices. Although *E*-values suggest robustness to unmeasured confounding, residual bias cannot be excluded. Finally, our analysis focused on physical outcomes; future work should examine how **β** shapes patient-reported outcomes, including satisfaction, decisional regret, and quality of life.

### β > Δα: BUT PATIENT CHOICE DOMINATES WHEN TREATMENTS ARE EQUAL

Many conditions carry clearly favorable treatment effects—settings in which patient choice plays little role because one option, one **α**, is clearly superior. In preference-sensitive settings such as SPORT, where treatment effects are similar, patient choice becomes proportionally more important. In Figure 1, the choice effect (panel c) exceeded the net treatment difference Δ**α c**(panel b) nearly 11-fold, indicating that preference-driven selection accounted for much of the divergence in nonrandomized comparisons. While generalizability beyond lumbar disc herniation requires further study, the framework likely extends to other preference-sensitive care decisions. In settings of clinical equipoise, decisions hinge less on biological efficacy and more on patient-specific utility—exactly the component quantified by **β**. Patient choice becomes, in the end, *dispositive*.

### CLINICAL INTERPRETATION OF β IN REAL_WORLD DECISION-MAKING

In preference-sensitive conditions, clinicians present options with comparable biological benefit. Outcomes then hinge on how patients interpret risk, trade short-term burden against long-term relief, and align treatment with their values and circumstances. Some accept operative risk to achieve faster symptom resolution; others prioritize avoiding surgery, even at the cost of slower recovery. These value-dependent choices shape adherence, engagement, and recovery trajectories. The parameter **β** captures this downstream effect of choice: it quantifies how selecting a preference-concordant treatment measurably alters physical outcomes, even when biological efficacy is similar.

This pattern is not unique to lumbar disc herniation. Many decisions—such as those involving prostate cancer, acute appendicitis, breast cancer, or atrial fibrillation—are characterized by clinical equipoise.^29,30,31,32^ In such circumstances, empirical evidence—and the AI systems trained on it—offers limited guidance in selecting a single superior option. Instead, patient values assume a central role not only in decision-making but in the outcomes that follow. The **α**/**β** framework makes explicit the distinction between biological efficacy and preference-mediated selection— two distinct, quantifiable drivers of outcome. Beyond its ethical imperative, patient choice can, in effect, be therapeutic in its own right.^33,34^

### MEDICAL LITERACY: THE “INFORMED” IN INFORMED CHOICE

The effect we call **β** is the causal footprint of patient choice, estimated alongside the biological treatment effect **α**; by definition, it does not require that choices be informed. Yet the magnitude and direction of **β** depend on whether patients can read diagnostic meaning, grasp risk, navigate options, and align decisions with their values—capacities that define medical literacy. Distinct from general education and from health literacy, which primarily supports prevention and longterm health maintenance, medical literacy governs escalation, option selection, adherence, and recovery. Gurel and Wilson formalize this as one of three disjoint education determinants acting on separate elements of a simple two-state public-health structure^35^. While not established here, it is reasonable to expect that informed choice—enabled by medical literacy—can amplify the **β** effect. Ongoing work outlines policy levers that build this substrate of comprehension, navigation, and preference-concordant decision-making^36^.

### WHAT AI CANNOT KNOW

Thus the same **β** that shows what patients must know—their values coupled to medical literacy— also shows what AI cannot: the lived, value-bound reasoning that no model can encode. Here the limits of current AI come into view: present-day systems face an epistemic boundary rooted in both their architecture and their training data. Large language models encode conditional probabilities from population-level evidence and excel at evidentiary synthesis and prediction. While **α** can be inferred from such evidence, typically derived from clinical trials, **β** remains the prerogative of individuals. Because the scientific literature informing medical AI assesses **α** almost exclusively, these systems remain confined to the biological treatment effect. In other words, AI is **α**-biased. Epistemically, the evidence base used to train AI systems—clinical trials, meta-analyses, and guidelines—focuses on average treatment effects (**α**) and rarely captures the deliberative processes that generate **β**.

The patient-choice effect we quantified arises from value functions that are counterfactual, heterogeneous, context-dependent, and normative. An LLM can correctly reproduce SPORT’s clinical equipoise, recognizing that surgery and nonoperative care yield similar outcomes. What it cannot access is the patient-specific utility function that determines whether surgical risk is acceptable for a given individual. That determination depends on **β**—a quantity not recoverable from **α**-biased outcome data. When treatment effects converge and **α** becomes non-determinative, **β** emerges as the dominant arbiter of outcome. AI systems optimized for predictive accuracy in such settings risk diminishing decision quality, because they conflate their strength—evidence synthesis—with their weakness—preference elicitation, a domain requiring human deliberation and judgment. Architecturally, current LLM-based systems lack mechanisms for structured, real-time preference elicitation and the contextual, normative reasoning needed to model patient-specific trade-offs; they are, in this sense, **β**-blind. As a result, AI-generated recommendations may misalign with patient goals, undermining shared decision-making and clinical outcomes. In principle, hybrid architectures that integrate explicit preference elicitation or utility modeling could mitigate this limitation, but such systems remain largely conceptual and are absent from the evidence base informing present-day clinical AI.

### IMPLICATIONS FOR AI AND MEDICINE

LLMs are ***smart***: adept at pattern recognition, evidence integration, and probabilistic reasoning. In preference-neutral conditions—where one treatment is clearly superior—LLMs may enhance care by synthesizing evidence, identifying subgroups, and predicting outcomes. They are not, however, ***wise***: they cannot align recommendations with patient-specific value systems or recognize when clinical equipoise demands preference elicitation rather than algorithmic advice. Recent evaluations of LLMs in clinical reasoning underscore these limitations, showing that systems trained solely on **α** cannot capture **β** and therefore cannot fully support decision-making in contexts where patient values are central^15^.

As noted, two challenges underlie this limitation: the **β**-blind architecture of AI and the **α**-biased data on which it is trained. Accordingly, two advances are needed. First, **improve the AI**: build hybrid systems fusing evidence synthesis with structured preference-elicitation tools, and add preference flags that detect equipoise and route decisions to shared deliberation. Second, **improve the data**: redesign clinical trials and care pathways to estimate **β** prospectively, elicit utilities and report **β**-identifying estimands alongside **α**. Without mechanisms to elicit and incorporate patient values, AI systems may misrepresent treatment efficacy, misrank options, or offer recommendations that conflict with patient goals. The **β**-blind spot we identified—a measurable and clinically meaningful causal component that current architectures cannot encode—marks a definitional limit of evidence-based AI in preference-sensitive care. Addressing these limitations requires hybrid architectures that integrate evidence synthesis with preference elicitation and a research infrastructure that captures the range of factors influencing clinical decisions.

## Data Availability

This study used no human-subject, clinical, or patient-level data. All information used is conceptual, theoretical, or derived from previously published sources. All computational spreadsheets and formulas used to generate illustrative estimates are available upon reasonable request.

## Acknowledgements

O. Gurel wishes to thank Dr. Xinlei Wang for the opportunity to present earlier versions of this work and Dr. Wei Jiang for helpful feedback. We are also grateful for thoughtful discussions with Dr. Zhezhen Jin, Dr. Amanda Evans, Dr. Peter Bonis, and Dr. Gabriela Mustata Wilson. Srujana Guggilla also supported analyses in the early stages of this work.

## SUPPLEMENTAL TABLE

**Table 1e.** Statistical Methods Comparison.

| **Table 1e - Statistical Methods Comparison** |  |  |  |  |  |
| --- | --- | --- | --- | --- | --- |
| **Method** | **Causal Inference** | **Handles Crossover** | **Selection Bias** | **NEJM Standard** | **Use** |
| **ITT** | Yes—randomization | No—diluted | No | Required baseline | Comparison |
| **IV/2SLS** | Yes—LATE | Yes—explicit | Via comparison | CONSORT guidelines | PRIMARY |
| **CACE** | Yes—compliers | Yes—isolates | Via comparison | CONSORT guidelines | CO-PRIMARY |
| **Observational** | No—confounded | N/A | Benchmark | Not alone | Bias quantification |
| **E-values** | N/A—sensitivity | N/A | Quantifies confounding | Recommended | ESSENTIAL |

## REFERENCES

1. Obermeyer Z, Emanuel EJ. Big data and machine learning. N Engl J Med 2016;375:1216–1219.

2. Topol EJ. High-performance medicine. Nat Med 2019;25:44–56.

3. Fahim YA, Hasani IW, Kabba S, Ragab WM. Artificial intelligence in healthcare and medicine: clinical applications, therapeutic advances, and future perspectives. Eur J Med Res 2025;30:848.

4. Andrès E, Escobar C, Doi K. Machine learning and artificial intelligence in clinical medicine— trends impact, and future directions. J Clin Med 2025;14:8137.

5. Wennberg JE. Preference-Sensitive Care: A Dartmouth Atlas Project Topic Brief. Lebanon (NH): The Dartmouth Institute for Health Policy and Clinical Practice; 2007 Jan 15. Available from: https://www.ncbi.nlm.nih.gov/books/NBK586631/

6. Wennberg JE. Unwarranted variations in healthcare delivery. BMJ 2002;325:961–964.

7. Wennberg JE. Time to tackle unwarranted variations in practice. BMJ 2011;342:d1513.

8. Weinstein JN, Tosteson TD, Lurie JD, et al. Surgical versus nonoperative treatment for lumbar disc herniation: the Spine Patient Outcomes Research Trial (SPORT): a randomized trial. JAMA 2006;296(18):2441–2450. doi:10.1001/jama.296.18.2441.

9. Weinstein JN, Lurie JD, Tosteson TD, et al. Surgical vs Nonoperative Treatment for Lumbar Disk Herniation: The Spine Patient Outcomes Research Trial (SPORT) Observational Cohort. JAMA 2006;296(20):2451–2459. doi:10.1001/jama.296.20.2451

10. Weinstein JN, Lurie JD, Tosteson TD, et al. Surgical versus nonoperative treatment for lumbar disc herniation: four-year results for the Spine Patient Outcomes Research Trial (SPORT). Spine 2008;33(25):2789–2800. doi:10.1097/BRS.0b013e31818ed8f4

11. McCormick PC. The Spine Patient Outcomes Research Trial results for lumbar disc herniation: a critical review. J Neurosurg Spine 2007;6:513–520. doi:10.3171/spi.2007.6.6.1

12. McCulloch P, Taylor I, Sasako M, Lovett B, Griffin D. Randomised trials in surgery: problems and possible solutions. BMJ 2002;324:1448–1451.

13. Elwyn G, Frosch D, Thomson R, et al. Shared decision making: a model for clinical practice. J Gen Intern Med 2012;27:1361–1367.

14. NICE Guideline NG197. Shared Decision Making. National Institute for Health and Care Excellence; 2021.

15. Yang M, Luo Y, He T, et al. Application of artificial intelligence to measure and predict patient values and preferences: a scoping review. npj Digit Med 2025;8:769. doi:10.1038/s41746-025-02156-2.

16. Aydin S, Karabacak M, Vlachos V, Margetis K. Navigating the potential and pitfalls of large language models in patient-centered medication guidance and self-decision support. Front Med 2025;12:1527864. doi:10.3389/fmed.2025.1527864.

17. Char DS, Shah NH, Magnus D. Implementing machine learning in health care — addressing ethical challenges. N Engl J Med 2018;378:981–3.

18. Imbens GW, Rubin DB. Causal Inference for Statistics, Social, and Biomedical Sciences: An Introduction. Cambridge University Press; 2015.

19. Lurie JD, Tosteson TD, Tosteson ANA, et al. Surgical vs nonoperative treatment for lumbar disc herniation: eight-year results for the Spine Patient Outcomes Research Trial. Spine 2014;39:3–16.

20. Mulley AG, Trimble C, Elwyn G. Patients’ preferences matter. BMJ 2012;345:e6572.

21. Barry MJ, Edgman-Levitan S. Shared decision making. N Engl J Med 2012;366:780–781.

22. Mannion AF, Junge A, Grob D, Dvorak J, Fairbank JCT. Development of a German version of the Oswestry Disability Index. Part 2: sensitivity to change after spinal surgery. Eur Spine J 2006;15(1):66–73. doi:10.1007/s00586-004-0816-z

23. Ogura K, Boland PJ, Bartelstein MK, Healey JH, Yakoub MA, Nikolic Z. Minimal clinically important differences in SF-36 global score: current value in orthopedic oncology. J Orthop Res 2020;38(12):2678–2684.

24. Fairbank JC, Pynsent PB. The Oswestry Disability Index. Spine 2000;25(22):2940–2952.

25. Ostelo RWJG, Deyo RA, Stratford P, et al. Interpreting change scores for pain and functional status in low back pain: towards international consensus regarding minimal clinically important change. Spine J 2008;8(6):968–974.

26. Solomito MJ, Kia C, Makanji H. The minimal clinically important difference for the Oswestry Disability Index substantially varies based on calculation method: implications to value-based care. Spine 2025;50(10):707–712. doi:10.1097/BRS.0000000000005074

27. Angrist JD, Imbens GW. Instrumental variables in randomized trials. N Engl J Med Evidence 2025;4:EVIDoa2400276.

28. Hernán MA, Robins JM. Instruments for causal inference. Epidemiology 2006;17:360–372.

29. Hamdy FC, Donovan JL, Lane JA, et al. 10-Year outcomes after monitoring, surgery, or radiotherapy for localized prostate cancer. N Engl J Med 2016;375:1415–24.

30. The CODA Collaborative. A randomized trial comparing antibiotics with appendectomy for appendicitis. N Engl J Med 2020;383:1907–19.

31. Hughes KS, Schnaper LA, Berry D, et al. Lumpectomy plus tamoxifen with or without irradiation in women 70 years of age or older with early breast cancer. N Engl J Med 2004;351:971–7.

32. January CT, Wann LS, Calkins H, et al. 2019 AHA/ACC/HRS focused update of the 2014 AHA/ACC/HRS guideline for the management of patients with atrial fibrillation: a report of the American College of Cardiology/American Heart Association Task Force on Clinical Practice Guidelines and the Heart Rhythm Society. Circulation 2019;140:e125–51.

33. Wasmann KA, Wijsman P, van Dieren S, Bemelman W, Buskens C. Partially randomised patient preference trials as an alternative design to randomised controlled trials: systematic review and meta-analyses. BMJ Open 2019;9(10):e031151. doi:10.1136/bmjopen-2019-031151

34. Delevry D, Le QA. Effect of treatment preference in randomized controlled trials: a systematic review and meta-analysis. The Patient 2019;12(6):593–609. doi:10.1007/s40271-019-00379-6

35. Gurel O, Mustata Wilson G. Education Is Three Determinants of Health—A Tripartite Classification. In Preparation; 2026.

36. Gurel O, Mustata Wilson G, Curley M, Weinstein JN. Public Health Policy Implications of Tripartite Education Determinants of Health. In Preparation; 2026.

